## Supplementary material for "Adjustment for Unmeasured Spatial Confounding in Settings of Continuous Exposure Conditional on the Binary Exposure Status: Conditional Generalized Propensity Score-Based Spatial Matching": Web Materials

**Web Table 1.** Data Sources and Variables Used in the Application Study

| **Type** | **Variable or data** | **Source** |
|---|---|---|
| Outcome | Census tract-level stroke prevalence (%) and coronary heart disease prevalence for the year 2018 | The Centers for Disease Control and Prevention (CDC) PLACES dataset |
| Exposure | Geocoded addresses for 59 petroleum refineries and petroleum production capacity for the year 2015–2017 | The United States Energy Information Administration |
| Confounder (Age) | the percentage of the population aged 18–19, 20–24, 25–44, 45–64, 65–84, and 85 years or older in the population aged 18 years or older | 5-year estimates from the American Community Survey (ACS, 2014–2018) |
| Confounder (Sex) | the percentage of males and females in the population aged 18 years or older | 5-year estimates from the American Community Survey (ACS, 2014–2018) |
| Confounder (Race/ethnicity) | The percentage of Hispanic, non-Hispanic white, non-Hispanic Black. | 5-year estimates from the American Community Survey (ACS, 2014–2018) |
| Confounder (SES) | Median household income, the percentages of the population who live under the federal poverty line or whose highest educational attainment is less than a high-school diploma | 5-year estimates from the American Community Survey (ACS, 2014–2018) |
| Confounder (smoking) | Self-reported current smoking for the year 2018 (%) | The Centers for Disease Control and Prevention (CDC) PLACES dataset |

**Web Figure 1. An Example of Distance-Matched Datasets With Estimated GPS. See Note.**

Suppose that one-to-n distance-matching gave us this dataset

| ID | $Z^b$ | $Z^c = w$ | Distance-matched stratum # | $Z^b_{cf}$ | $Z^c_{cf}$ | $GPS \equiv f(Z^c_{cf} = w \mid \boldsymbol{C})$ | Duplicated after the distance-matching with replacement? |
|---|---|---|---|---|---|---|---|
| 1 | 1 | 100 | 1 | 1 | 100 | $f(Z^c = 100 \mid \boldsymbol{C}) = P(Z^b = 1 \mid \boldsymbol{C}) \times f(Z^c = 100 \mid \boldsymbol{C}, Z^b = 1) = 0.45$ | No |
| 2 | 0 | 0 | 1 | 1 | 100 | $f(Z^c_{cf} = 100 \mid \boldsymbol{C}) = P(Z^b_{cf} = 1 \mid \boldsymbol{C}) \times f(Z^c_{cf} = 100 \mid \boldsymbol{C}, Z^b_{cf} = 1) = 0.20$ | No |
| 3 | 0 | 0 | 1 | 1 | 100 | $f(Z^c_{cf} = 100 \mid \boldsymbol{C}) = P(Z^b_{cf} = 1 \mid \boldsymbol{C}) \times f(Z^c_{cf} = 100 \mid \boldsymbol{C}, Z^b_{cf} = 1) = 0.41$ | Yes |
| 4 | 1 | 50 | 2 | 1 | 50 | $f(Z^c = 50 \mid \boldsymbol{C}) = P(Z^b = 1 \mid \boldsymbol{C}) \times f(Z^c = 50 \mid \boldsymbol{C}, Z^b = 1) = 0.34$ | No |
| 3 | 0 | 0 | 2 | 1 | 50 | $f(Z^c_{cf} = 50 \mid \boldsymbol{C}) = P(Z^b_{cf} = 1 \mid \boldsymbol{C}) \times f(Z^c_{cf} = 50 \mid \boldsymbol{C}, Z^b_{cf} = 1) = 0.33$ | Yes |
| 5 | 0 | 0 | 2 | 1 | 50 | $f(Z^c_{cf} = 50 \mid \boldsymbol{C}) = P(Z^b_{cf} = 1 \mid \boldsymbol{C}) \times f(Z^c_{cf} = 50 \mid \boldsymbol{C}, Z^b_{cf} = 1) = 0.32$ | No |
| … | … | … | … | | | … | … |

After the distance-matching, one-to-one nearest neighbor matching *without* replacement by GPS will give us…

| ID | $Z^b$ | $Z^c = w$ | Distance-matched stratum # | $Z^b_{cf}$ | $Z^c_{cf}$ | $GPS \equiv f(Z^c_{cf} = w \mid \boldsymbol{C})$ |
|---|---|---|---|---|---|---|
| 1 | 1 | 100 | 1 | 1 | 100 | $f(Z^c = 100 \mid \boldsymbol{C}) = P(Z^b = 1 \mid \boldsymbol{C}) \times f(Z^c = 100 \mid \boldsymbol{C}, Z^b = 1) = 0.45$ |
| 3 | 0 | 0 | 1 | 1 | 100 | $f(Z^c_{cf} = 100 \mid \boldsymbol{C}) = P(Z^b_{cf} = 1 \mid \boldsymbol{C}) \times f(Z^c_{cf} = 100 \mid \boldsymbol{C}, Z^b_{cf} = 1) = 0.41$ |
| 4 | 1 | 50 | 2 | 1 | 50 | $f(Z^c = 50 \mid \boldsymbol{C}) = P(Z^b = 1 \mid \boldsymbol{C}) \times f(Z^c = 50 \mid \boldsymbol{C}, Z^b = 1) = 0.34$ |
| 5 | 0 | 0 | 2 | 1 | 50 | $f(Z^c_{cf} = 50 \mid \boldsymbol{C}) = P(Z^b_{cf} = 1 \mid \boldsymbol{C}) \times f(Z^c_{cf} = 50 \mid \boldsymbol{C}, Z^b_{cf} = 1) = 0.32$ |
| … | … | … | … | | | … |

One-to-one nearest neighbor matching *with* replacement by GPS will give us…

| ID | $Z^b$ | $Z^c = w$ | Distance-matched stratum # | $Z^b_{cf}$ | $Z^c_{cf}$ | $GPS \equiv f(Z^c_{cf} = w \mid \boldsymbol{C})$ | Duplicated after the distance-matching with replacement? |
|---|---|---|---|---|---|---|---|
| 1 | 1 | 100 | 1 | 1 | 100 | $f(Z^c = 100 \mid \boldsymbol{C}) = P(Z^b = 1 \mid \boldsymbol{C}) \times f(Z^c = 100 \mid \boldsymbol{C}, Z^b = 1) = 0.45$ | No |
| 3 | 0 | 0 | 1 | 1 | 100 | $f(Z^c_{cf} = 100 \mid \boldsymbol{C}) = P(Z^b_{cf} = 1 \mid \boldsymbol{C}) \times f(Z^c_{cf} = 100 \mid \boldsymbol{C}, Z^b_{cf} = 1) = 0.41$ | Yes |

| | | | | | | | |
|---|---|---|---|---|---|---|---|
| 4 | 1 | 50 | 2 | 1 | 50 | $f(Z^c = 50 \mid \boldsymbol{C}) =$ $P(Z^b = 1 \mid \boldsymbol{C}) \times f(Z^c = 50 \mid \boldsymbol{C}, Z^b = 1) = 0.34$ | No |
| 3 | 0 | 0 | 2 | 1 | 50 | $f\left(Z_{cf}^c = 50 \middle\| \boldsymbol{C}\right) =$ $P\left(Z_{cf}^b = 1 \middle\| \boldsymbol{C}\right) \times f\left(Z_{cf}^c = 50 \middle\| \boldsymbol{C}, Z_{cf}^b = 1\right) = 0.33$ | Yes |
| … | … | … | … | | | … | … |

Note: ID 3 (unexposed unit; in red) is matched to ID 1 (exposed unit; in blue) and to ID 4 (exposed unit; in blue) by one-to-n distance-matching with replacement. GPS of ID 3 in the distance-matched stratum #1 is different than GPS of ID 3 in the distance-matched stratum #2 because $Z_{cf}^c$ is different per $w$ in each stratum. In the GPS-matching step, one-to-one nearest neighbor matching without replacement will give us a pair of ID 1 and ID 3 at the distance-matched stratum #1 and will give us a pair of ID 4 and ID 5 at the distance-matched stratum #2. For the latter, although GPS of ID 3 (0.33) is closer to GPS of ID 4 (0.34) than GPS of ID 5 (0.32), ID 4 is matched to ID 5 because ID 3 is already matched to ID 1 at the distance-matched stratum #1. In contrast, one-to-one nearest neighbor matching with replacement will match ID 4 to ID 3.

**Web Figure 2. Spatial Patterns of *U* with Nine Pairs of the Smoothness and Range Parameters of the Matérn Covariance Function.**

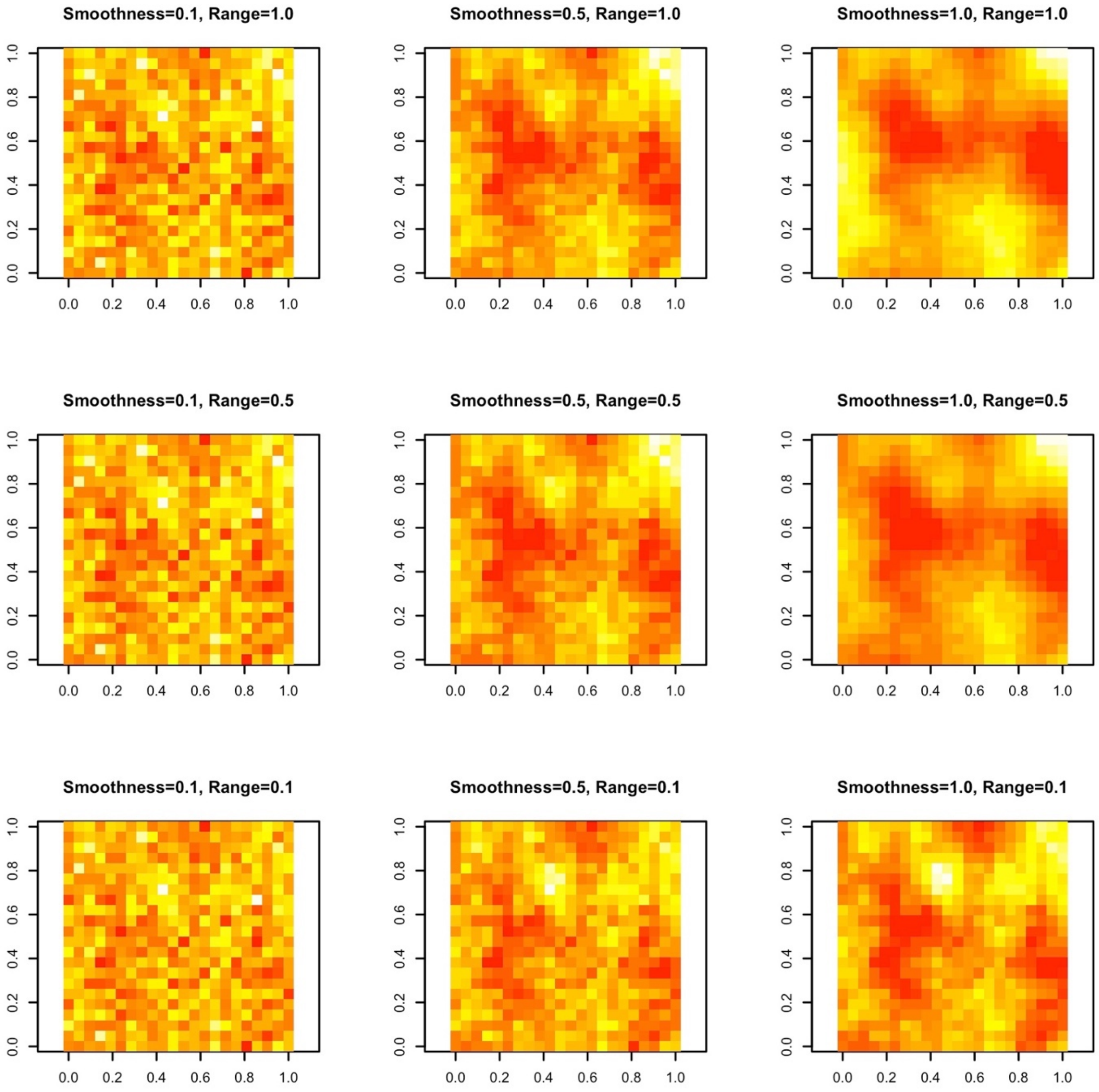

### 1. Detailed Methods in the Simulation Study

#### Data generating process

For every pair of two spatial parameters (smoothness parameter, $k$; range parameter, $\pi$) we simulated 200 datasets for which a total of 484 (22×22) fixed locations were simulated as follows.

A. One unmeasured confounder, *U*, was generated from a Gaussian Process with Mat´ern correlation function. *U* was normalized to have mean 0 and variance 1.

B. Three observed confounders, $X_1$, $X_2$, and $X_3$ were independently generated from the standardized normal distribution (mean=0; variance=1).

C. A binary exposure, $Z_b$, was generated from a logistic model

$$logit(P(Z_b)) = -3 + 1X_1 + 1.4X_2 + 0.8X_3 + 1.3U$$

This generating model yields approximately 15% of the observational units exposed.

D. A continuous exposure, $Z_c$ was generated from a linear regression model

$$Z_c = 50 + 2X_1 + 4X_2 + 3.5X_3 + 6U + \epsilon \quad \epsilon \sim N(0,5^2)$$

E. An outcome, *Y*, was generated from a Poisson model

$$\log(Y) = 0.03Z_c + 0.15X_1 + 0.23X_2 + 0.31X_3 + U$$

#### Matching process and estimation of ATT

A. One-to-n distance-matching: we matched exposed units to unexposed units if their spatial distance is lower than equal to 0.1 degree. This matching was done with replacement.

B. We fit a PS model with $X_1$, $X_2$, and $X_3$, and coordinates. We fit a generalized additive model (GAM) with the logistic function, binomial distribution, and spatial smoother using coordinates (a thin-plate spline) or used the eXtreme gradient boosting algorithm (XGBoost). For XGBoost, the objective function was a logistic regression for binary classification. The evaluation metric was log-loss. To select hyperparameters (e.g., maximum depth of a tree, maximum number of trees, the learning rate), we used grid-search with 10-fold cross-validation. For the methods with $U$ known, we fit a logistic regression model including $X_1$, $X_2$, $X_3$, and $U$ instead.

C. We fit a CGPS model with $X_1$, $X_2$, and $X_3$, and coordinates using the exposed subset of the dataset. We fit a GAM with the identity function, gaussian distribution, and spatial smoother using coordinates (a thin-plate spline). For the methods with $U$ known, we fit a linear regression model including $X_1$, $X_2$, $X_3$, and $U$ instead.

D. We estimated GPS as described in the main text.

E. GPS-matching: For every pair of exposed units and unexposed units matched by the distance, we matched exposed units and unexposed units by GPS. We used one-to-one nearest-neighbor matching with/without replacement and one-to-one nearest-neighbor caliper matching with/without replacement.

F. We generated 500 bootstrapping samples from the distance- and GPS-matched dataset.

G. We fit a disease model to estimate ATT using each of the bootstrapped samples. We used *R* package *gnm* to fit conditional Poisson regression models (with eliminate option for the matched strata). The output of this analysis is equivalent to that of Poisson regression analysis with dummy variables for matched strata added.

H. We obtained mean of coefficient estimates and mean of standard error estimates over the bootstrapped samples.

### 2. Detailed Methods in the Application Study

A. One-to-n distance matching: We matched exposed units to unexposed units if their spatial distance is lower than equal to 0.1 degree. This matching was done with replacement.

B. We fit a PS model with pre-selected measured potential confounders (Table S1), and coordinates. Selection of confounder variables was made considering standardized mean differences. We fit a generalized additive model (GAM) with the logistic function, binomial distribution, and spatial smoother using coordinates (a thin-plate spline) or used the eXtreme gradient boosting algorithm (XGBoost). For XGBoost, the objective function was a logistic regression for binary classification. The evaluation metric was log-loss. To select hyperparameters (e.g., maximum depth of a tree, maximum number of trees, the learning rate), we used grid-search with 10-fold cross-validation.

C. We fit a CGPS model with pre-selected measured potential confounders and coordinates using the exposed subset of the dataset. For exposed units, $X_{ct}$ was log-transformed. Selection of confounder variables was made considering correlations between measured potential confounders and $X_{ct}$ in the exposed subset. We used GAM with the identity function, gaussian distribution, and spatial smoother using coordinates (a thin-plate spline).

D. We estimated GPS as described in the main text.

E. GPS-matching: For every pair of exposed units and unexposed units matched by the distance, we matched exposed units and unexposed units by GPS. We used one-to-one nearest-neighbor matching with/without replacement and one-to-one nearest-neighbor caliper matching with/without replacement.

F. We checked balance in measured potential confounders using standardized mean differences.

G. If covariates were imbalanced, we repeated steps B-F by adding different confounder variables to PS and CGPS models. When they were balanced, we generated 500 bootstrapping samples using the distance- and GPS-matched dataset.

H. We fit a disease model to estimate ATT using each of the bootstrapped samples. We used *R* package *gnm* to fit conditional Poisson regression models (with eliminate option for the matched strata). The output of this analysis is equivalent to that of Poisson regression analysis with dummy variables for matched strata added.

I. We obtained mean of coefficient estimates and mean of standard error estimates over the bootstrapped samples.

**Web Figure 3. Bias by Traditional Regression With/Without *U*, Naïve IPW-GAM, and Naïve IPW-XGBoost.**

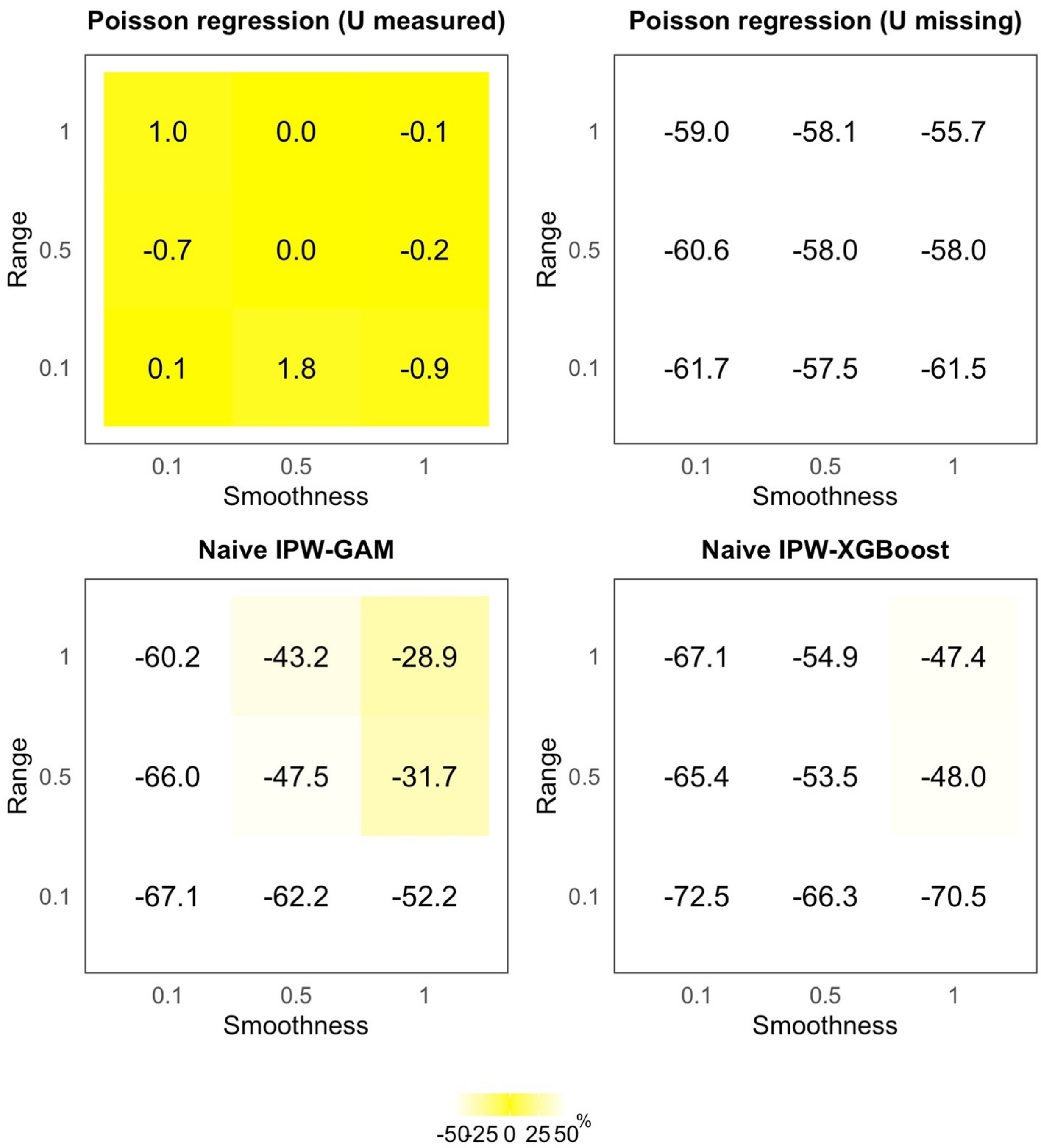

**Web Figure 4. Nominal Coverage of 95% Confidence Intervals by Traditional Regression With/Without U, Naïve IPW-GAM, and Naïve IPW-XGBoost.**

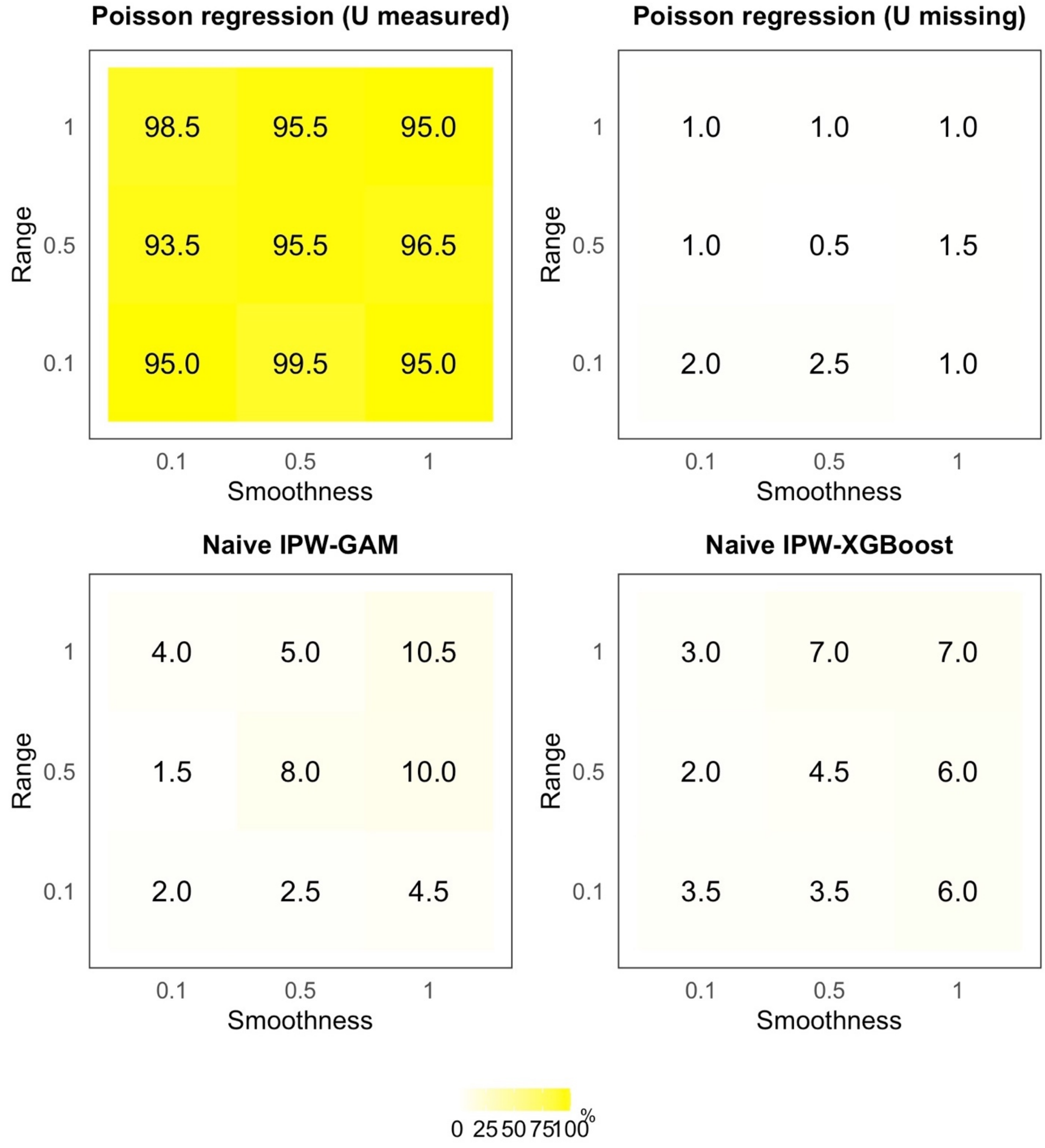

**Web Figure 5. Bias by CGPSsm Methods Without Replacement.**

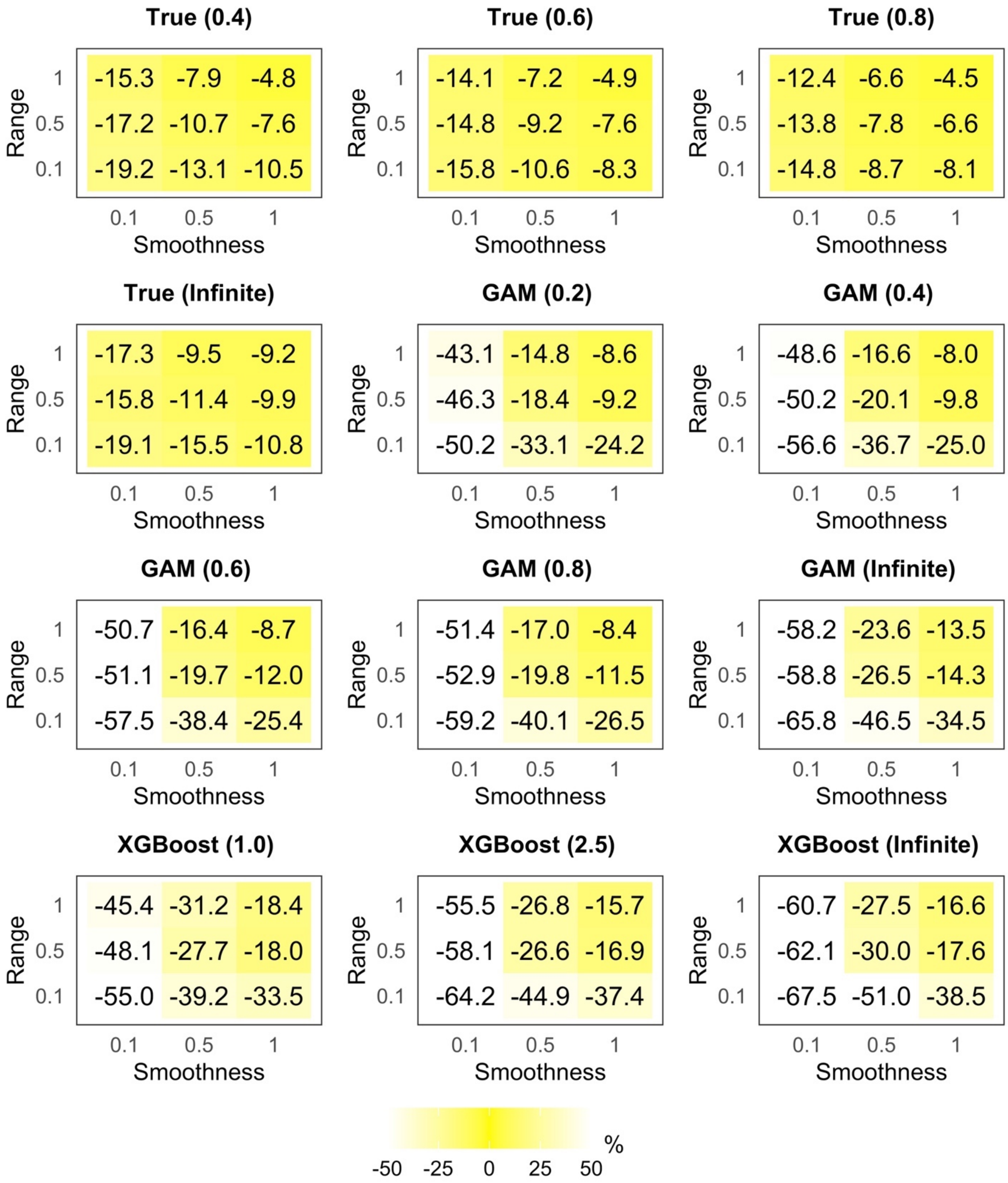

**Web Figure 6. Root Mean Squared Error by CGPSsm Methods Without Replacement**

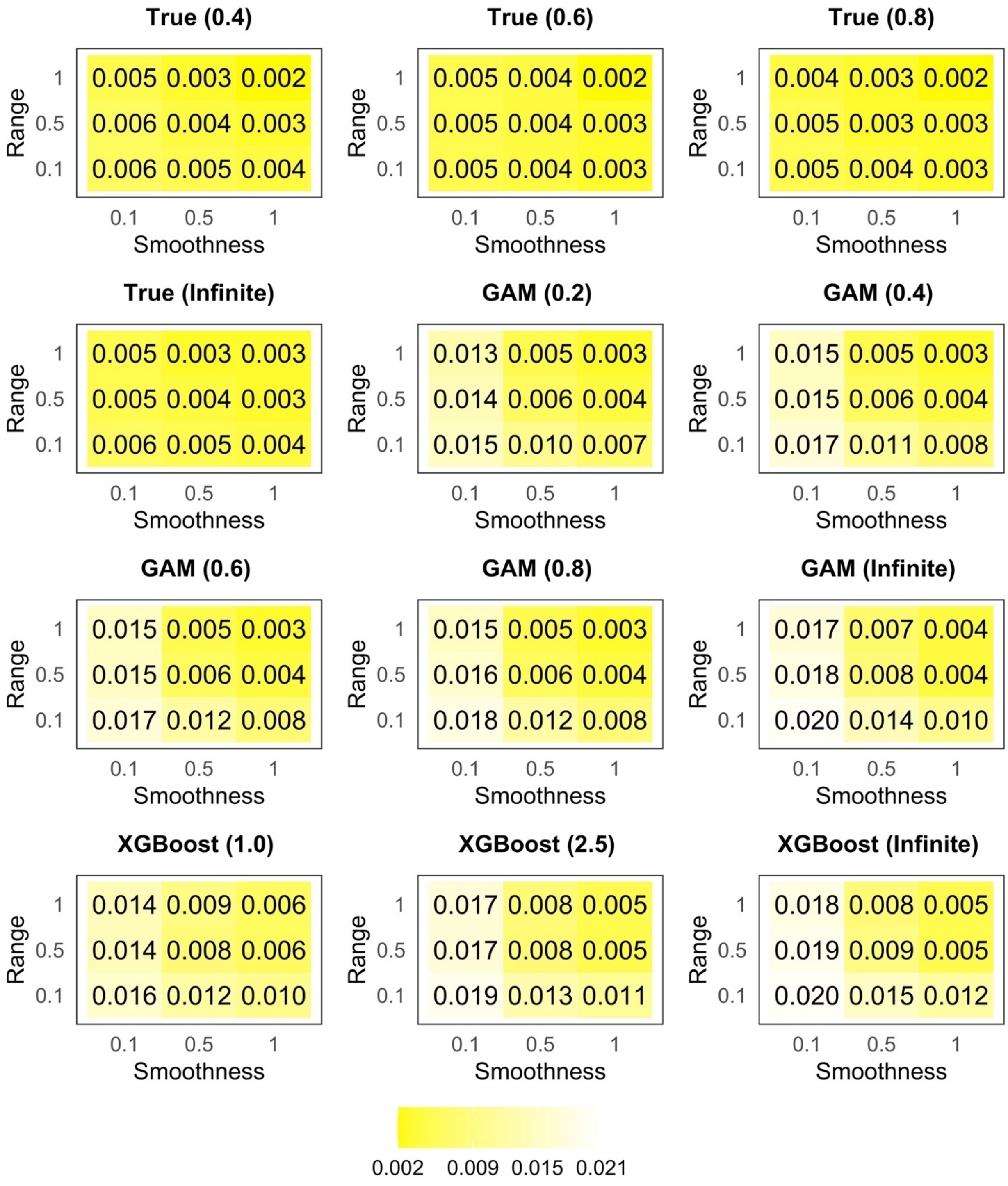

**Web Figure 7. Nominal Coverage of 95% Confidence Intervals by CGPSsm Methods Without Replacement.**

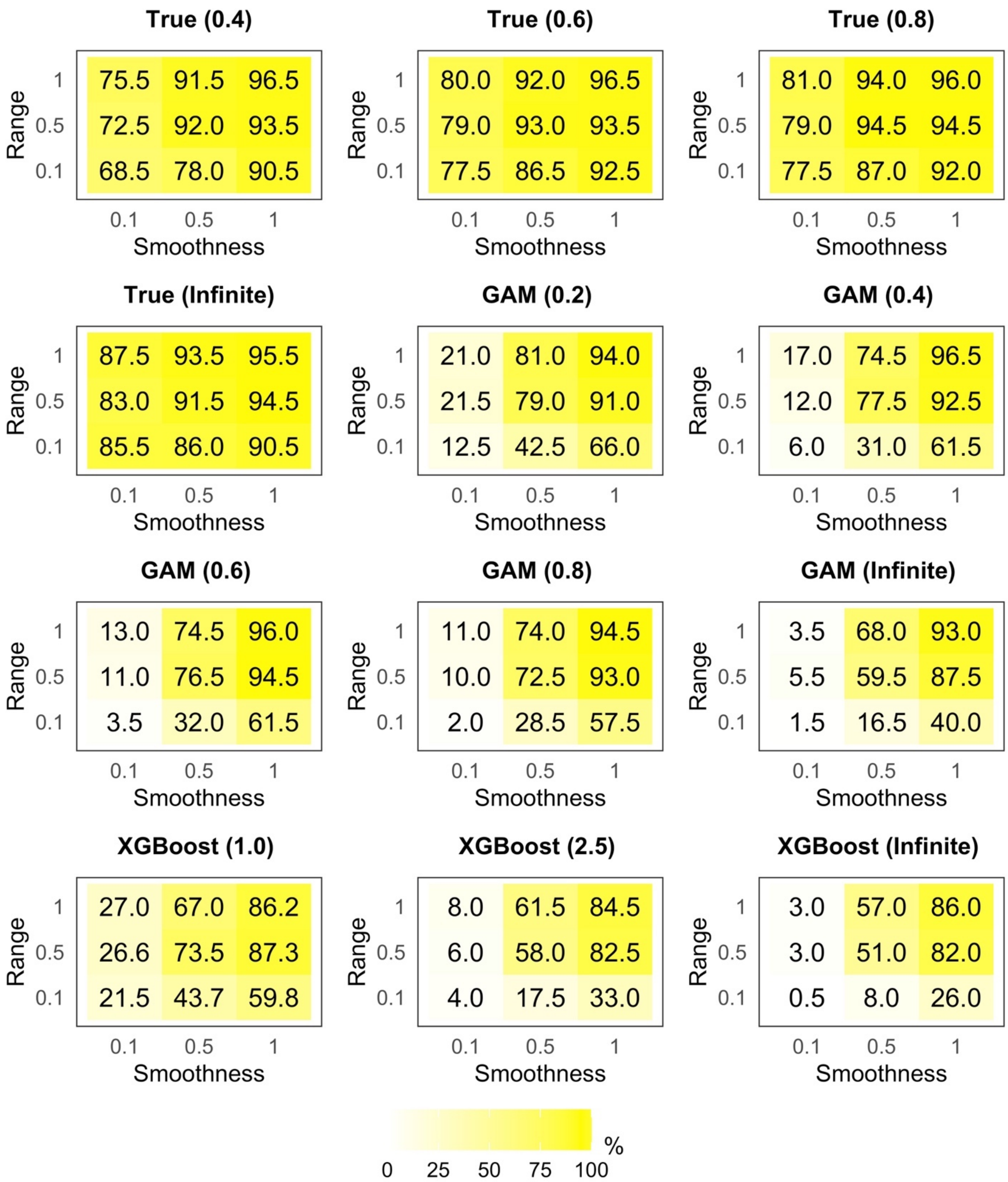

**Web Figure 8. Percentage of Exposed Units Matched to Unexposed Units by CGPSsm Methods Without Replacement.**

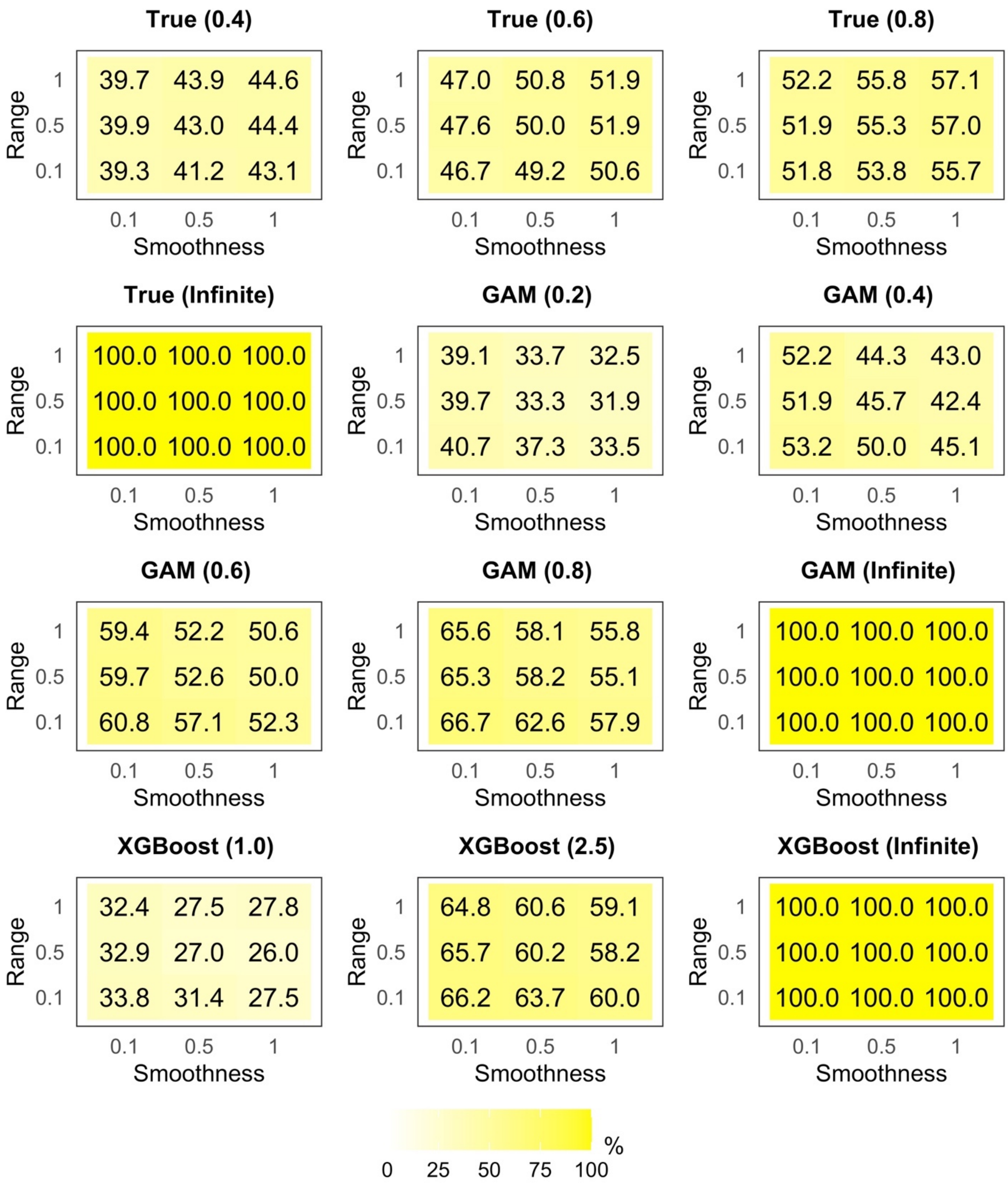

**Web Figure 9. Covariate Balance Before and After CGPSsm Without Replacement.**

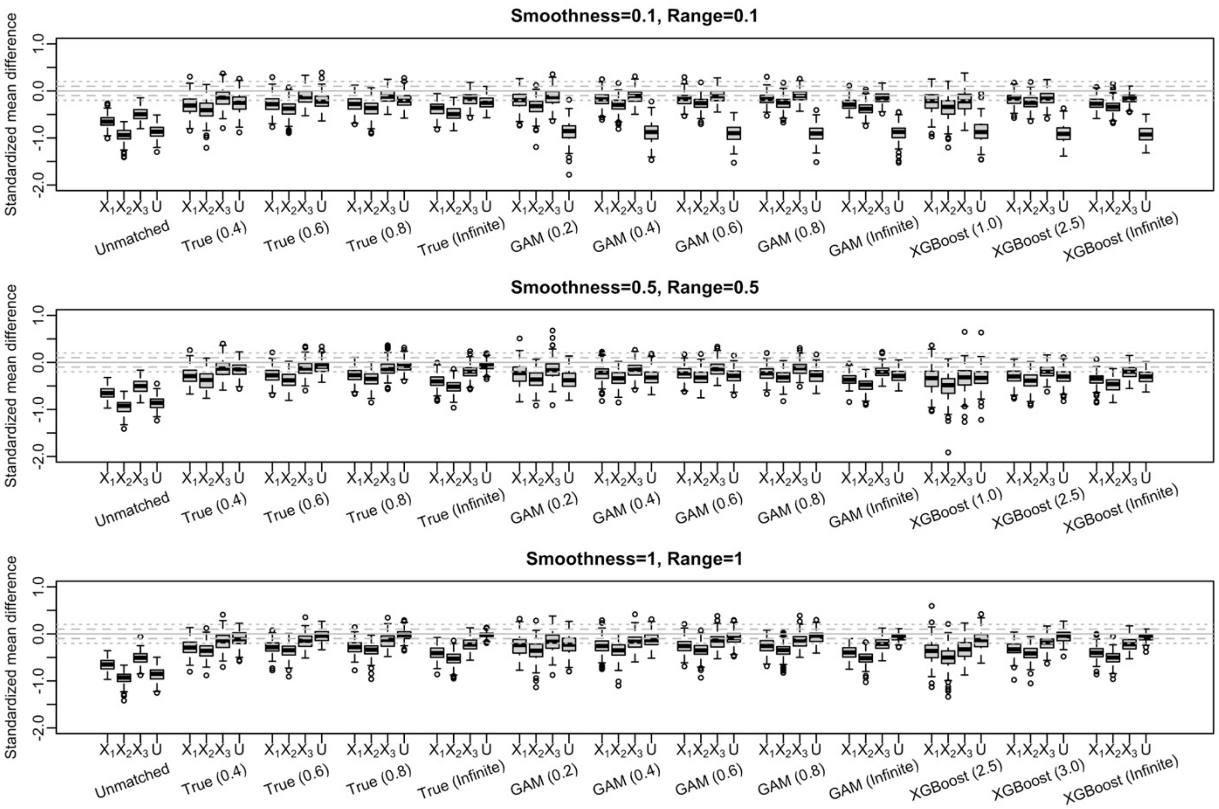

**Web Figure 10. Covariate Balance Before/After CGPSsm in The Motivating Example.**

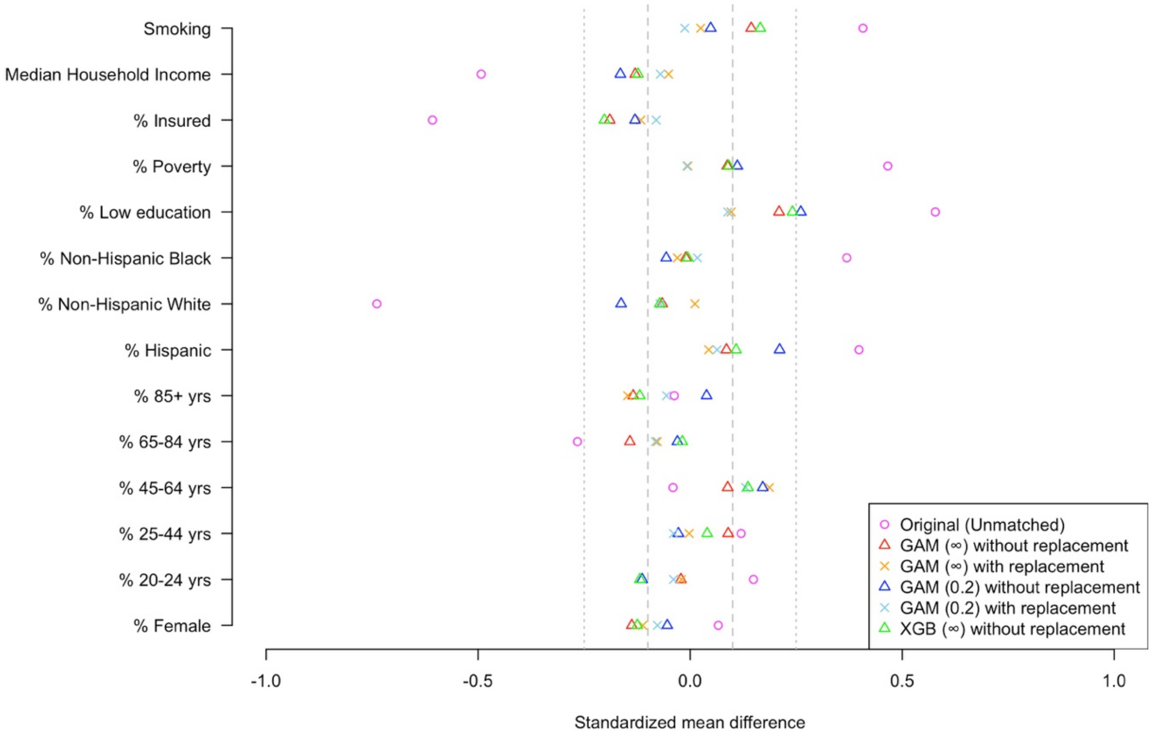

Note: The number in brackets indicate $cw$ in one-to-one nearest neighbor caliper matching; $cw = \infty$ indicates one-to-one nearest neighbor matching; Grey dashed lines indicate ±0.1. Grey dotted lines indicate ±0.25.
